# Validity and Reliability of the CVI Range for Clinical Research (CVI Range-CR): Baseline and One-Year Results

**DOI:** 10.1101/2025.10.17.25338230

**Authors:** Melinda Y. Chang, Mark W. Reid, Christine Roman-Lantzy, Dilshad Contractor, Katherine Granger, Sharon H. O’Neil, Francesca Crozier-Fitzgerald, Martha Veto, Mark S. Borchert

## Abstract

**Purpose:** Cerebral/cortical visual impairment (CVI) is a leading cause of visual impairment in children, and significantly impacts both visual function and functional vision (the ability to use vision during real-life activities). We developed the CVI Range for Clinical Research (CVI-CR), a functional vision assessment that can be incorporated in CVI clinical trials. Herein we evaluate the psychometric properties, inter- and intra-rater reliability, and validity of the CVI Range-CR.

**Design:** Prospective, longitudinal cohort study

**Participants:** 40 children with CVI

**Methods:** Participants underwent CVI Range-CR assessments at baseline and one-year follow-up. All assessments were recorded. A trained in-person examiner scored in-person assessments; two expert remote graders scored recorded assessments. Visual acuity was evaluated by a pediatric neuro-ophthalmologist using the six-level visual behavior scale (VBS).

**Main Outcome Measures:** CVI Range-CR scores (ranging from 0 to 10) using the Across-CVI Characteristics (Rating 1) and Within-CVI Characteristics (Rating 2) methods of scoring were compared among graders using the intraclass correlation coefficient (ICC) to assess inter-rater reliability. A subset of videos was scored twice by the same remote examiner to assess intra-rater reliability using the ICC. CVI Range-CR scores were compared to VBS scores (Spearman’s correlation coefficient, ⍴) to evaluate validity. Cronbach’s ɑ was used to assess internal consistency of the 10 items on the Within-CVI Characteristics scale.

**Results:** The CVI Range-CR demonstrated high internal consistency (Cronbach’s ɑ=0.96), excellent intra-rater reliability (ICC=0.88-0.98), good-to-excellent inter-rater reliability (ICC=0.74-0.88), and strong correlation to clinical assessment of visual behavior as measured by VBS scores (Spearman’s ⍴=−0.76 to −0.86, p<0.0001). It was also sensitive to change; there were small but significant improvements in both Ratings 1 and 2 over one year (Rating 1 average: +0.35, p=0.002; Rating 2 average: +0.31, p=0.001).

**Conclusions:** The CVI Range-CR is a reliable and valid measure of functional vision in children with CVI. Because intra-rater reliability exceeds inter-rater reliability, longitudinal studies are recommended to use consistent graders over time. This could be accomplished by remote grading of recorded assessments via a centralized reading center.

## Introduction

Cerebral/cortical visual impairment (CVI) is a leading cause of visual impairment in children in the United States and other developed countries, and is increasing in developing countries as a result of improved survival of children born with serious neurological conditions.^1–8^ CVI refers to “a spectrum of visual impairments caused by an underlying brain abnormality that affects the development of visual processing pathways and is characterized by deficits in visual function and functional vision.”^9^ Visual function refers to measures that are assessed in the eye clinic, such as acuity and contrast sensitivity. Functional vision refers to how a person uses vision in everyday activities.^10^ In individuals with CVI, there may be a disconnect between visual function and functional vision such that their ability to use vision during real-life activities is worse than expected based on visual function measured in the eye clinic.^11, 12^ This may be related to higher-level visual processing deficits that are not assessed during standard eye exams,^13^ which may significantly impair quality of life.^14^ Thus, evaluation of functional vision is crucial in children with CVI and must be performed using specialized techniques.

Methods to evaluate functional vision include standardized history-taking questionnaires^15–17^ and functional vision assessments administered by trained personnel.^18, 19^ Tests of higher-order visual processing such as standardized psychometric assessments of visual perception^20, 21^ and certain eye tracking protocols,^22–24^ especially those that incorporate virtual reality,^23^ may also provide clues about a child’s ability to perform everyday activities. The most widely used functional vision assessment in children with CVI is the CVI Range, developed by Dr. Roman-Lantzy.^18^ The CVI Range has been used by teachers for the visually impaired, occupational therapists, and other low vision specialists to guide individualized interventions to improve a child’s functioning at school and home. For this reason, the CVI Range has often been performed in a non-standardized manner to facilitate personalized recommendations. However, a standardized functional vision assessment is necessary to serve as an outcome measure in clinical trials. This is critical to ensure that medical treatments proposed for CVI impact a child’s ability to function at home and school, not just in the eye clinic.

We developed the CVI Range for Clinical Research (CVI Range-CR),^25^ a standardized version of the CVI Range that can be incorporated into clinical research studies.^18^ The CVI Range-CR is administered by a trained examiner who conducts a family interview and direct assessment of the child with CVI, while observing how the child interacts with visual stimuli strategically placed in a standard configuration around the assessment room.^25^ The assessment is scored using two scales, the Across-CVI characteristics (Rating 1) and Within-CVI characteristics (Rating 2) methods, with scores that range from 0 to 10. The Within-CVI characteristics method is divided into 10 domains, based on 10 CVI characteristics (Figure 1A). CVI Range-CR scores are divided into three progressively increasing phases of functional vision in CVI (Figure 1C). The CVI Range-CR can be video recorded and scored remotely, thereby facilitating scoring of assessments at a centralized reading center in future clinical trials.

**Figure 1.**
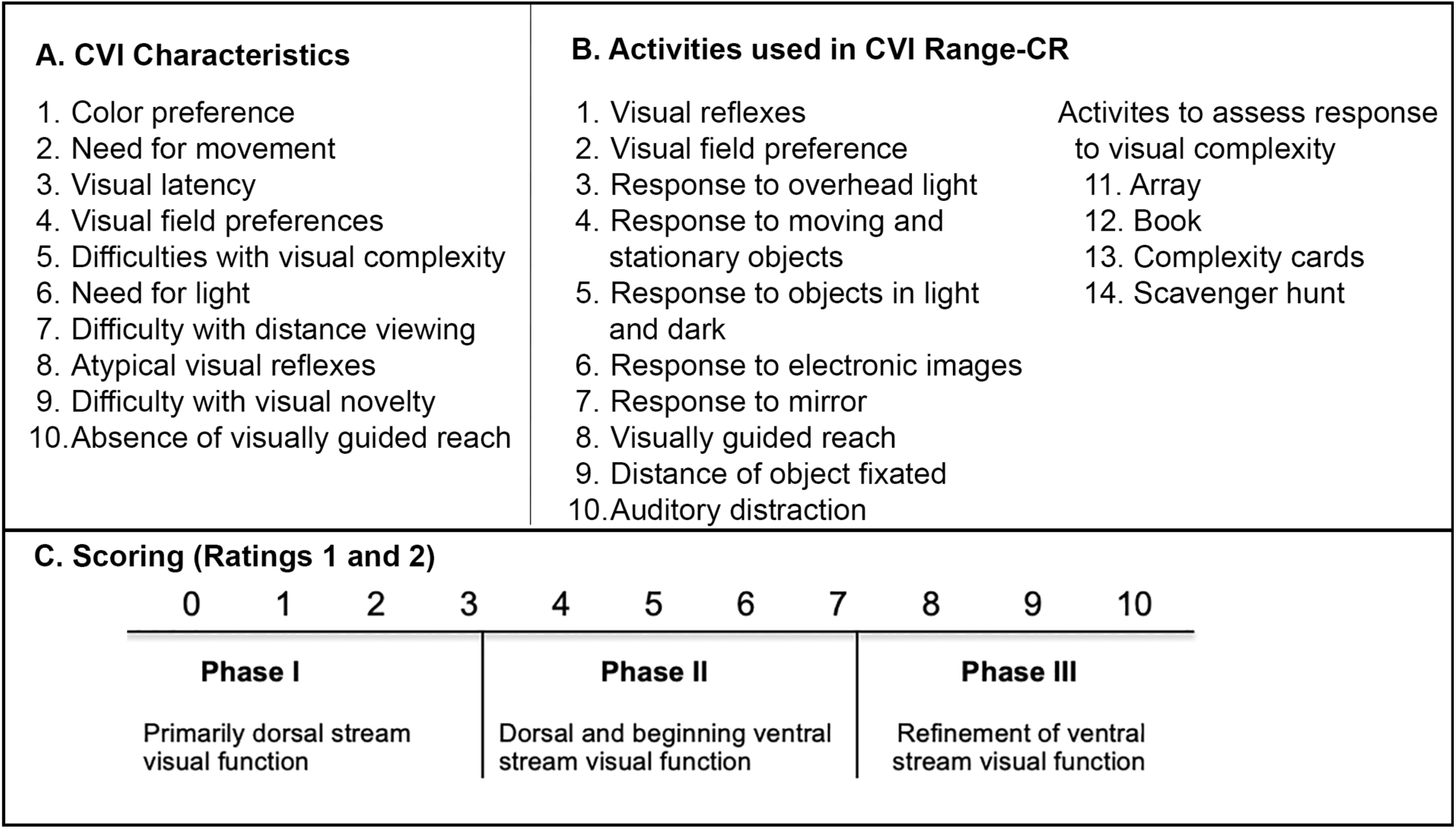
Summary of CVI Range-CR. A) Ten CVI characteristics evaluated; B) 14 activities performed by the examiner; C) Range of scores and corresponding phases.

The purpose of this study was to assess the internal consistency, intra-rater reliability, inter-rater reliability, and validity of the CVI Range-CR functional vision assessment.

## Methods

This study was approved by the Children’s Hospital Los Angeles Institutional Review Board (IRB) and adhered to the tenets of the Declaration of Helsinki and the US Health Insurance Portability and Accountability Act of 1996. Informed consent was obtained from the parent or legal guardian of all participants.

We prospectively recruited children between 12 months and 10 years of age with a diagnosis of CVI from our pediatric neuro-ophthalmology clinic. CVI was diagnosed in children with decreased visual acuity for age with a normal eye exam, or worse than expected visual acuity based on the degree of ocular pathology, in the context of a structural or functional neurologic disorder affecting the post-geniculate visual pathways in the brain. Although CVI can occur in children with normal visual acuity and higher-order visual processing deficits,^13, 16, 26^ we did not include these patients in our study because the overwhelming majority of CVI patients in our clinic have moderately to severely reduced visual acuity. We excluded patients with ophthalmoplegia or any limitation of ocular ductions that would preclude assessment of visual latency, which is an important characteristic of CVI. However, patients with strabismus and nystagmus with full ocular ductions were included. We also excluded families who were not fluent in English, since the interview was conducted in English.

### Overview of study visits

Participants underwent two study visits, at baseline and 1 year follow-up. Each study visit was coordinated with a clinic visit with a pediatric neuro-ophthalmologist, who performed a complete eye exam including assessment of visual acuity. At each study visit, parents completed questionnaires and a CVI Range-CR assessment was conducted by a neuropsychologist.

### Visual Behavior Scale

Because our CVI participants were unable to perform optotype acuity testing, we used the six-level visual behavior scale (VBS)^27^ to grade visual acuity based on the ability to fixate and follow objects of various sizes at various distances (Supplemental Table 1). Level 6 indicates the worst visual acuity (no response to light), and level 1 is the best (ability to accurately fixate and follow a 1 inch toy at 1 foot). The pediatric neuro-ophthalmologist who performed VBS grading was masked to the CVI Range-CR score.

### Parent questionnaires

At baseline and follow-up visits, parents completed the baseline and follow-up questionnaires, respectively (Supplemental Forms). These surveys provided information on demographics, medical history, and developmental assessments and services.

Additionally, at both visits parents completed the Vineland Adaptive Behavior Scales, 3^rd^ edition (VABS-III), a standardized psychometric measure of adaptive functioning.^28^ The VABS-III provides an overall Adaptive Behavior Composite (ABC) score, as well as scores on several domains (and subdomains): Communication (Receptive, Expressive, Written), Daily Living Skills (Personal, Domestic, Community), and Socialization (Interpersonal Relationships, Play and Leisure Time, Coping Skills).

### CVI Range-CR

The CVI Range-CR functional vision assessment was performed at both visits as previously described.^25^ Briefly, a trained neuropsychologist (SHO), who was masked to VBS scores, served as the in-person examiner and grader. First, she conducted a family interview using a list of 24 standard questions.^25^ Next, she performed direct assessment of the child’s functional vision by guiding the child through a series of up to 14 activities (Figure 1B) designed to assess 10 CVI characteristics (Figure 1A). A standard set of materials, which are commercially available and can be obtained online, were used.^25^ Throughout the assessment, the examiner observed whether the child spontaneously interacted with items placed around the assessment room, without directing their attention to these objects. The total time for each assessment was approximately 1 hour.

#### CVI Range-CR recording

During each visit, a research coordinator or assistant recorded the CVI Range-CR assessment using two iPads placed orthogonally to one another. The recordings were later synced and combined into a single video using iMovie (Apple Inc, Cupertino, CA). During this process, the video editor also removed any instances of personal health information (PHI) being inadvertently shared, such as the examiner using the child’s name.

#### CVI Range-CR scoring

The in-person examiner (SHO) and two expert remote graders (FCF, MV) scored the CVI Range-CR assessments using the Across-CVI Characteristics (Rating 1) and Within-CVI Characteristics (Rating 2) scales (Supplemental Forms). In the Across-CVI characteristics method of scoring, graders assessed visual functioning across five blocks of behaviors representing increasing levels of functioning. In the Within-CVI characteristics method of scoring, graders assigned scores for each of the 10 CVI characteristics ranging from 0 to 1 (in increments of 0.25), and the total score was the sum of domain scores. Higher scores indicate more advanced functional vision. We input both the Across- and Within-CVI Characteristics scoring sheets into a REDCap database,^29^ which was programmed to calculate both scores.

The in-person examiner scored all assessments based on observations made during the visit. The remote graders scored based on video recordings. At baseline, one remote grader was assigned to grade each video (assignments were alternated). At 1 year, both remote graders scored all videos. This resulted in one grader masked and one grader unmasked to the baseline visit for each participant at the 1 year follow-up visit.

Additionally, approximately 1 year after baseline visits were complete, we randomly chose 7 baseline videos and requested them to be regraded by the same remote grader that was initially assigned. This enabled us to calculate intra-rater reliability.

#### Examiner drift

In order to correct any drift in performance of the CVI Range-CR assessment by the in-person examiner, video recordings were reviewed periodically by the author of the original CVI Range functional vision assessment (CRL). After every six recordings, two videos were reviewed. Feedback was provided to the in-person examiner if drift was noted.

### Statistical analysis

Demographic and clinical characteristics were summarized using descriptive statistics (mean±standard deviation, or count and percentage). Differences in these characteristics between participants who completed one-year follow-up visits and those who did not were evaluated using Mann Whitney U-tests or Fisher’s Exact tests, as appropriate.

Intraclass Correlation Coefficients (ICC) were used to evaluate both intra- and inter-rater reliability, where values closer to 1 suggest stronger agreement. For intra-rater reliability, absolute agreement values with 95% confidence intervals (CI) are reported. For inter-rater reliability, both absolute and relative agreement, where individual differences among graders are controlled, are reported. Linear mixed effects (LME) models were used to calculate ICC values, allowing for inclusion of data from participants who were not evaluated by all three graders.

The internal consistency of the 10-item Within-CVI Characteristics (Rating 2) score was evaluated using inter-item correlations, Cronbach’s α, and McDonald’s ω, a generalized form of Cronbach’s α that allows for uneven factor loadings. A confirmatory factor analysis was also conducted to evaluate the structure of Rating 2, which was expected to reflect a single construct.

Correlations between CVI Range-CR scores and other clinical measures were calculated as Spearman rank-order correlations using LME models to account for clustered measurements per participant and grader. Changes in clinical outcomes from baseline to follow-up visit were also examined using LME models, which allow for inclusion of repeated measures of participant data (i.e., baseline and follow-up visits) and evaluation of fixed differences in grader behavior.

All analyses were conducted using Stata/SE 14.2 (College Station, TX, USA). For all analyses, p-values less than 0.05 were considered significant.

### Sample size calculation

The sample size for this study was selected in advance based on the number of patients required to detect an ICC with sufficient precision – at least good agreement (between 0.60 and 0.74), where the lower bound of the CI around the estimate exceeds 0.50. Based on these criteria, a sample of 36 children with CVI would be required to detect an ICC as low as 0.68, assuming error rates α=0.05 and β=0.20 (80% power). To account for up to 20% attrition, we aimed to recruit 43 participants. In addition, we calculated that remote graders would each need to re-evaluate at least 7 of their own videos to produce estimates of self-agreement with sufficient precision (ICC>0.80).

## Results

### Participants

We recruited 40 participants who completed CVI Range-CR and VBS assessments at baseline. Of these, 36 (90%) completed the VABS-III questionnaire, and 31 (78%) completed the baseline questionnaire.

One-year follow-up CVI Range-CR and VBS assessments were conducted on 34 participants (85%); the remaining six were lost to follow-up. Of the 34 retained participants, 32 (94%) completed the one-year VABS-III questionnaire, and 23 (68%) completed the follow-up study questionnaire.

The demographic and clinical characteristics of the full cohort at baseline are shown in Table 1. The mean age was 4.3±2.5 years. The mean VBS score was 2.8±1.1 on the six-level VBS scale, indicating the average participant could redirect gaze to a face or large toy, but could not accurately fixate and follow a 6-inch toy at 1 foot. The mean CVI Range-CR Rating 1 score was 3.5±1.7, and the Rating 2 score was 3.4±1.6, indicating that on average participants were in early phase 2 when starting this study. There was no significant difference in demographic or clinical characteristics between the participants who completed the 1-year follow-up visit (n=34) and those who did not (n=6).

**Table 1.**
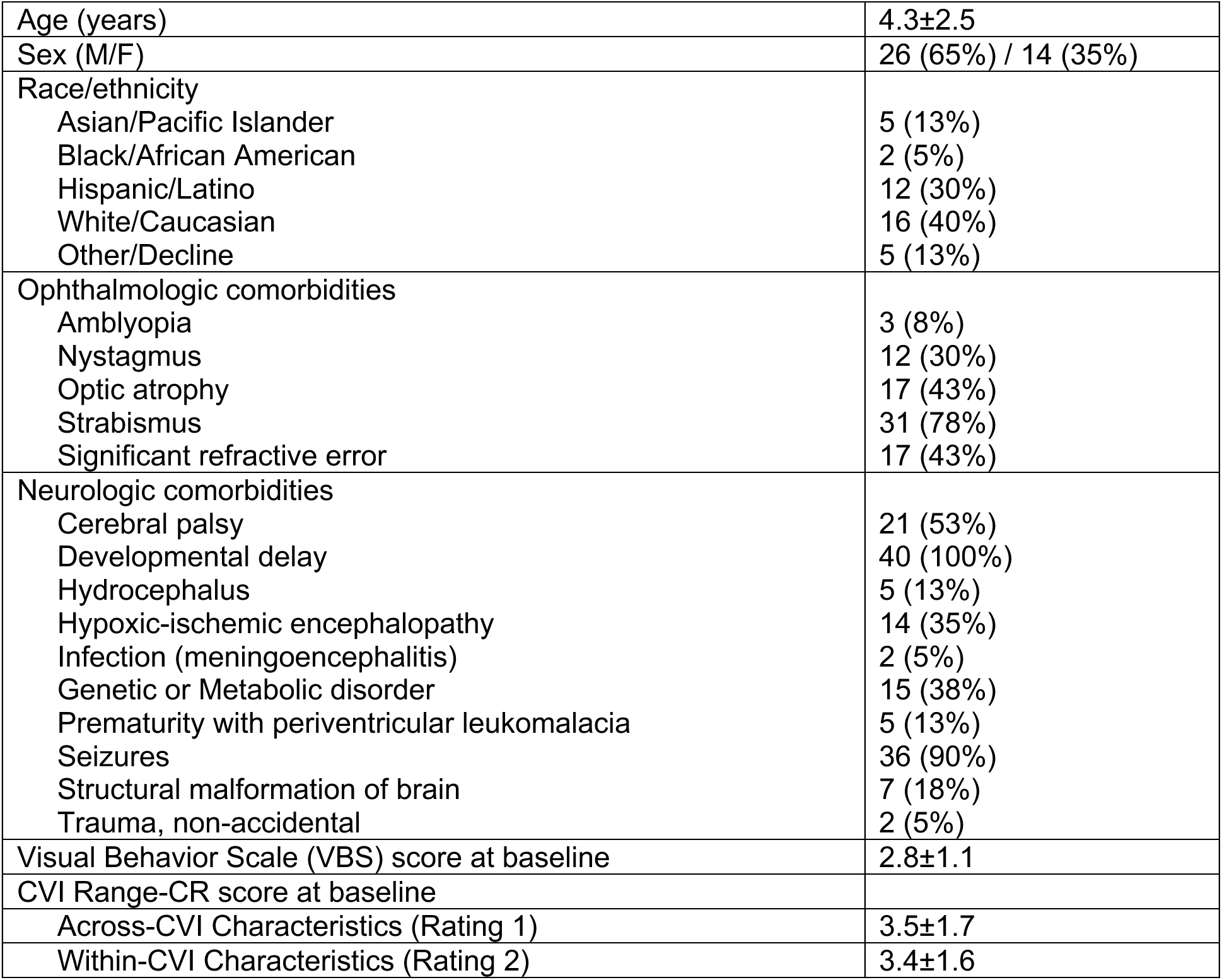
Clinical and demographic characteristics of 40 children with cerebral/cortical visual impairment (CVI). Means and standard deviations are reported for continuous and ordinal data.

|  |  |
| --- | --- |
| Age (years) | 4.3±2.5 |
| Sex (M/F) | 26 (65%) / 14 (35%) |
| Race/ethnicity |  |
| Asian/Pacific Islander | 5 (13%) |
| Black/African American | 2 (5%) |
| Hispanic/Latino | 12 (30%) |
| White/Caucasian | 16 (40%) |
| Other/Decline | 5 (13%) |
| Ophthalmologic comorbidities |  |
| Amblyopia | 3 (8%) |
| Nystagmus | 12 (30%) |
| Optic atrophy | 17 (43%) |
| Strabismus | 31 (78%) |
| Significant refractive error | 17 (43%) |
| Neurologic comorbidities |  |
| Cerebral palsy | 21 (53%) |
| Developmental delay | 40 (100%) |
| Hydrocephalus | 5 (13%) |
| Hypoxic-ischemic encephalopathy | 14 (35%) |
| Infection (meningoencephalitis) | 2 (5%) |
| Genetic or Metabolic disorder | 15 (38%) |
| Prematurity with periventricular leukomalacia | 5 (13%) |
| Seizures | 36 (90%) |
| Structural malformation of brain | 7 (18%) |
| Trauma, non-accidental | 2 (5%) |
| Visual Behavior Scale (VBS) score at baseline | 2.8±1.1 |
| CVI Range-CR score at baseline |  |
| Across-CVI Characteristics (Rating 1) | 3.5±1.7 |
| Within-CVI Characteristics (Rating 2) | 3.4±1.6 |

### Inter-rater reliability

ICC values reflecting inter-rater reliability are shown in Table 2. Overall, raters showed good-to-excellent absolute agreement for both Rating 1 (absolute ICC=0.76, 95% CI: 0.61-0.91) and Rating 2 (absolute ICC=0.74, 95% CI: 0.50-0.98). When variations in individual rater behavior were accounted for, agreement was excellent while confidence intervals narrowed (Rating 1: relative ICC=0.84, 95% CI: 0.75-0.90; Rating 2: relative ICC=0.88, 95% CI: 0.81-0.92). Agreement for each domain of Rating 2 varied depending on content, ranging in absolute agreement from ICC=0.48 to 0.73, and relative agreement from ICC=0.56 to 0.77. Differences between absolute and relative agreement indicate the presence of fixed variations in behavior of the raters, suggesting that certain raters consistently scored higher than others. This difference was most apparent in the Need for Light domain.

**Table 2.** Inter-rater reliability of 3 graders (one in-person, two remote) for 40 CVI Range-CR assessments at baseline and 34 assessments at 1 year follow-up.

|  | **Absolute Agreement** |  | **Relative Agreement** |  |
| --- | --- | --- | --- | --- |
|  | **ICC** | **95% CI** | **ICC** | **95% CI** |
| Rating 1 – Across-CVI characteristics | 0.76 | 0.61 – 0.91 | 0.84 | 0.75 – 0.90 |
| Rating 2 – Within-CVI characteristics | 0.74 | 0.50 – 0.98 | 0.88 | 0.81 – 0.92 |
| Rating 2 Domains |  |  |  |  |
| 1. Color Preference | 0.64 | 0.47 – 0.81 | 0.71 | 0.59 – 0.81 |
| 2. Need for Movement | 0.58 | 0.39 – 0.77 | 0.66 | 0.53 – 0.77 |
| 3. Visual Latency | 0.63 | 0.40 – 0.85 | 0.74 | 0.63 – 0.83 |
| 4. Visual Field Preferences | 0.51 | 0.31 – 0.71 | 0.60 | 0.45 – 0.72 |
| 5. Difficulties with Visual Complexity | 0.54 | 0.31 – 0.76 | 0.65 | 0.51 – 0.76 |
| 6. Need for Light | 0.48 | 0.24 – 0.72 | 0.62 | 0.48 – 0.74 |
| 7. Difficulty with Distance Viewing | 0.66 | 0.50 – 0.82 | 0.72 | 0.60 – 0.81 |
| 8. Atypical Visual Reflexes | 0.59 | 0.41 – 0.77 | 0.67 | 0.53 – 0.78 |
| 9. Difficulty with Visual Novelty | 0.46 | 0.24 – 0.68 | 0.56 | 0.42 – 0.70 |
| 10. Absence of Visually Guided Reach | 0.73 | 0.60 – 0.86 | 0.77 | 0.68 – 0.85 |

ICC values of inter-rater reliability that consider only baseline or follow-up assessments showed similar findings to the combined analysis (Supplemental Tables 2 and 3).

### Intra-rater reliability

ICC values reflecting intra-rater reliability are shown in Table 3. Overall, intra-rater reliability was excellent (Rating 1: ICC=0.88 to 0.92; Rating 2: ICC=0.94 to 0.98), indicating high reliability for repeated assessments of a patient by the same rater. Rating 2 domains with the lowest intra-rater reliability were Visual Field Preferences (ICC=0.63 to 0.73), Need for Light (ICC=0.68 to 0.71) Atypical Visual Reflexes (ICC=0.43 to 0.88), and Difficulty with Visual Novelty (ICC=0.51 to 1.00). Self-agreement varied by domain, suggesting that each rater may struggle to interpret some individual characteristics, but their overall scores are robust against these minor variations in subdomains.

**Table 3.** Intra-rater reliability of two remote graders for seven CVI Range-CR assessments.

|  | **Grader 1** |  | **Grader 2** |  |
| --- | --- | --- | --- | --- |
|  | **ICC** | **95% CI** | **ICC** | **95% CI** |
| Rating 1 – Across-CVI characteristics | 0.92 | 0.63 – 0.98 | 0.88 | 0.49 – 0.98 |
| Rating 2 – Within-CVI characteristics | 0.98 | 0.88 – 1.00 | 0.94 | 0.73 – 0.99 |
| Rating 2 Domains |  |  |  |  |
| 1. Color Preference | 0.80 | 0.27 – 0.96 | 0.68 | 0.00 – 0.93 |
| 2. Need for Movement | 0.73 | 0.11 – 0.95 | 0.82 | 0.34 – 0.97 |
| 3. Visual Latency | 0.83 | 0.36 – 0.97 | 0.88 | 0.51 – 0.98 |
| 4. Visual Field Preferences | 0.73 | 0.11 – 0.95 | 0.63 | -0.07 – 0.92 |
| 5. Difficulties with Visual Complexity | 0.73 | 0.11 – 0.95 | 0.77 | 0.21 – 0.96 |
| 6. Need for Light | 0.71 | 0.07 – 0.94 | 0.68 | 0.00 – 0.93 |
| 7. Difficulty with Distance Viewing | 0.73 | 0.11 – 0.95 | 0.87 | 0.46 – 0.97 |
| 8. Atypical Visual Reflexes | 0.88 | 0.51 – 0.98 | 0.43 | -0.34 – 0.87 |
| 9. Difficulty with Visual Novelty | 1.00 | – | 0.51 | -0.24 – 0.89 |
| 10. Absence of Visually Guided Reach | 0.88 | 0.52 – 0.98 | 0.94 | 0.73 – 0.99 |

### Internal Consistency (Within-CVI Characteristics scale, Rating 2)

The structure of Rating 2 of the CVI Range-CR, which evaluates functioning across 10 domains corresponding to 10 CVI characteristics, allows for analysis to determine how strongly the 10 characteristics are related to one another (Table 4). Item-test correlations ranged from 0.69 to 0.91, with the lowest correlation found with the Atypical Visual Reflexes domain. Even though scores on this domain did not covary as strongly with those on other domains, the overall Cronbach’s ɑ internal consistency score of the measure was unchanged when the item was removed. Both Cronbach’s α and McDonald’s ω were 0.96, indicating strong internal consistency.

**Table 4.** Inter-item correlations and internal consistency on the CVI Range-CR Within-CVI Characteristics scale (Rating 2).

| Domain | N Obs. | Item-Test Correlation | Item-Rest Correlation | Correlation w/out Item | Alpha $\alpha$ w/out Item |
| --- | --- | --- | --- | --- | --- |
| Color Preference | 182 | 0.88 | 0.84 | 0.69 | 0.95 |
| Need for Movement | 182 | 0.84 | 0.80 | 0.69 | 0.95 |
| Visual Latency | 182 | 0.91 | 0.88 | 0.68 | 0.95 |
| Visual Field Preferences | 182 | 0.84 | 0.80 | 0.70 | 0.95 |
| Difficulty with Visual Complexity | 182 | 0.89 | 0.86 | 0.68 | 0.95 |
| Need for Light | 182 | 0.85 | 0.81 | 0.69 | 0.95 |
| Difficulty with Distance Viewing | 182 | 0.87 | 0.84 | 0.69 | 0.95 |
| Atypical Visual Reflexes | 182 | 0.69 | 0.61 | 0.73 | 0.96 |
| Difficulty with Visual Novelty | 182 | 0.86 | 0.82 | 0.69 | 0.95 |
| Absence of Guided Reach | 174 | 0.89 | 0.86 | 0.68 | 0.95 |
| **Overall Scale Consistency** |  |  |  |  |  |
| **Cronbach's $\alpha$:** |  |  |  |  | **0.96** |
| **McDonald's $\omega$:** |  |  |  |  | **0.96** |

N. Obs. = number of observations, w/out = without

Confirmatory factor analysis verified a single-factor structure for Rating 2, with a root mean square error of approximation (RMSEA) of 0.045 and coefficient of determination (R^2^) of 0.965, indicating excellent model fit for all 10 items.

### Convergent Validity

*VBS Visual Acuity.* The correlation between CVI Range-CR scores and pediatric neuro-ophthalmologic assessment of visual acuity using the VBS is provided in Table 5. All three graders showed good to excellent correlation with VBS scores on both Ratings 1 and 2 (Spearman’s correlation coefficient −0.76 to −0.85, p<0.0001). When comparing the average CVI Range-CR scores of the 3 graders to the VBS scores, the Spearman’s correlation coefficient was −0.85 for Rating 1 and −0.86 for Rating 2 (p<0.0001 for both).

**Table 5.**
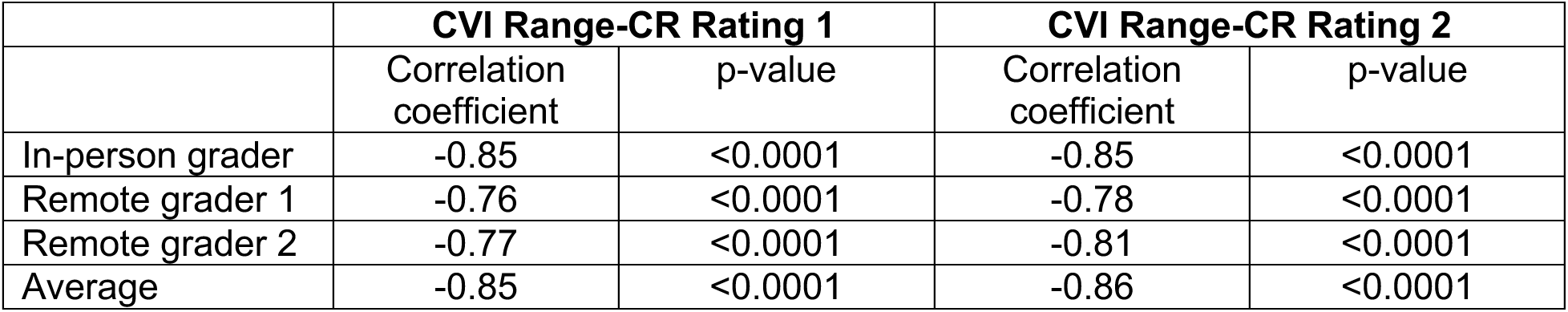
Correlation between CVI Range-CR and Visual Behavior Scale (VBS) scores, using a mixed model to account for assessments at both baseline and 1 year follow-up visits.

Similar findings were seen when only baseline or follow-up CVI Range-CR scores were correlated with VBS scores (Supplemental Tables 4 and 5).

*VABS-III Adaptive Behavior*. The correlation between CVI Range-CR scores (averaged among the 3 graders) and VABS-III scores, including both baseline and follow-up visits, is shown in Table 6. There was statistically significant correlation between CVI Range-CR and VABS-III scores on the Adaptive Behavior Composite and most domains and subdomains. However, the strength of the correlations ranged from weak to moderate. The VABS-III domain that correlated most strongly with CVI Range-CR scores was Motor Skills (Spearman’s correlation coefficient 0.527 to 0.529, p<0.001).

**Table 6.** Correlation between CVI Range-CR and Vineland Adaptive Behavior Scales, 3^rd^ edition (VABS-III) scores, using a mixed model to include both baseline and 1 year follow-up assessments. The average CVI Range-CR scores from multiple graders were used for this analysis.

|  | CVI Range-CR Rating 1 |  | CVI Range-CR Rating 2 |  |
| --- | --- | --- | --- | --- |
|  | Correlation coefficient | p-value | Correlation coefficient | p-value |
| Adaptive Behavior Composite (ABC) | 0.384 | 0.002 | 0.373 | 0.002 |
| Communication |  |  |  |  |
| Domain | 0.429 | 0.001 | 0.418 | 0.001 |
| Subdomains |  |  |  |  |
| Receptive | 0.351 | 0.010 | 0.343 | 0.010 |
| Expressive | 0.245 | 0.069 | 0.279 | 0.030 |
| Written | 0.174 | 0.226 | 0.231 | 0.087 |
| Daily Living Skills |  |  |  |  |
| Domain | 0.421 | 0.001 | 0.403 | 0.001 |
| Subdomains |  |  |  |  |
| Personal | 0.354 | 0.005 | 0.377 | 0.002 |
| Domestic | 0.297 | 0.032 | 0.303 | 0.021 |
| Community | 0.162 | 0.221 | 0.235 | 0.054 |
| Socialization |  |  |  |  |
| Domain | 0.357 | 0.004 | 0.372 | 0.002 |
| Subdomains |  |  |  |  |
| Interpersonal Relationships | 0.453 | <0.001 | 0.439 | <0.001 |
| Play and Leisure | 0.435 | <0.001 | 0.457 | <0.001 |
| Coping Skills | 0.342 | 0.011 | 0.374 | 0.004 |
| Motor Skills |  |  |  |  |
| Domain | 0.529 | <0.001 | 0.527 | <0.001 |
| Subdomains |  |  |  |  |
| Gross Motor | 0.421 | 0.001 | 0.425 | 0.001 |
| Fine Motor | 0.377 | 0.001 | 0.342 | 0.003 |

The results of correlation analysis between CVI Range-CR scores and VABS-III scores reported separately at baseline and follow-up visits are shown in Supplemental Tables 6 and 7, respectively.

### Change in Scores over Follow-Up

Figure 2 illustrates the change in CVI Range-CR scores per grader over the one-year follow-up period. Both Ratings 1 and 2 showed small but significant improvements (Rating 1 average: +0.35, p=0.002; Rating 2 average: +0.31, p=0.001). Figure 2 also shows the scores for each grader at baseline and follow-up. In general, the differences in scores between the 3 raters remained similar at baseline and follow-up visits, especially for Rating 2. This is consistent with our finding that ICC scores of relative agreement were higher than scores of absolute agreement, indicating that fixed grader differences partly accounted for variability in scores.

**Figure 2.**
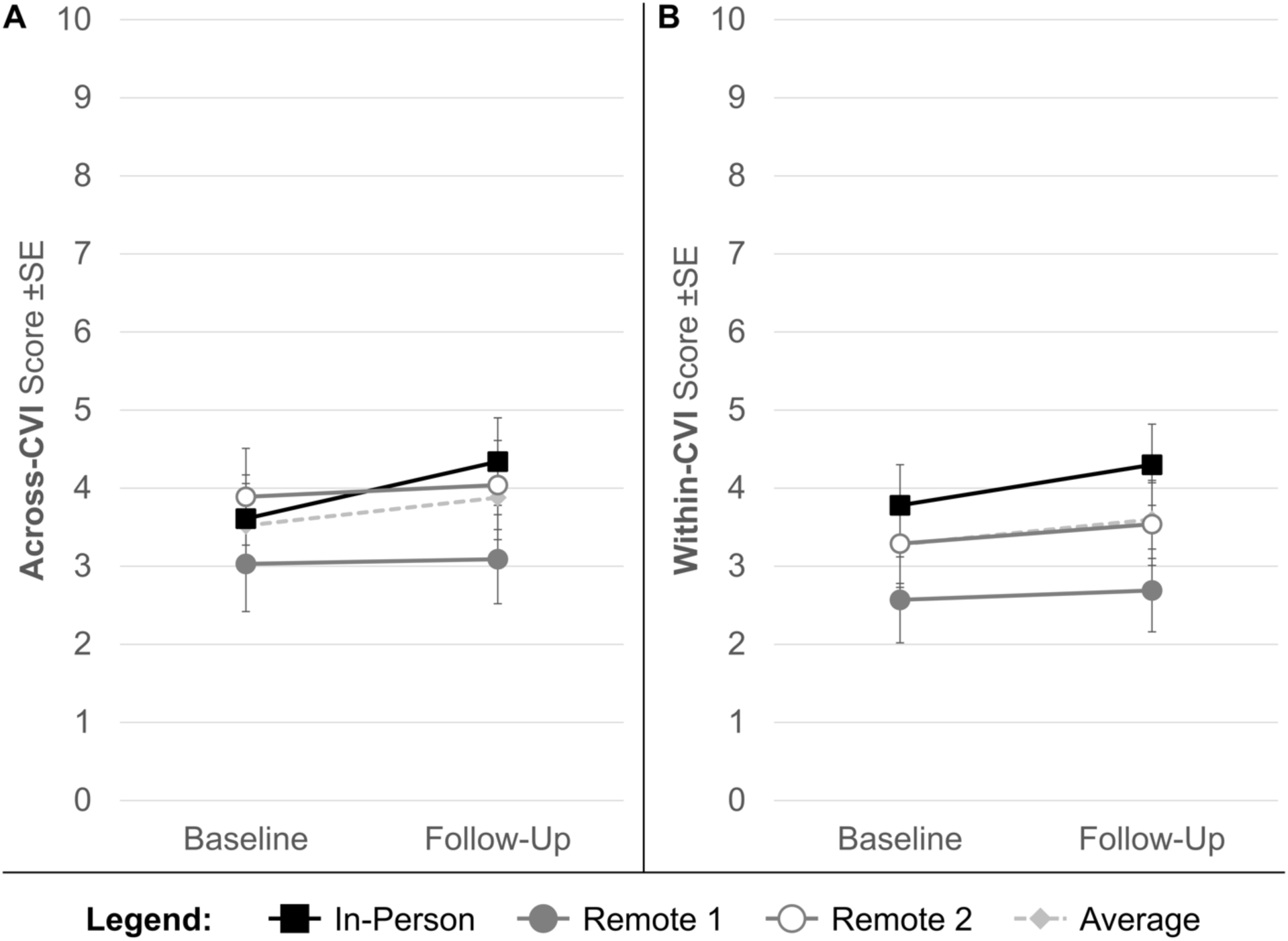
Change in CVI Range-CR scores over 1 year, by grader. A) Across-CVI Characteristics score (Rating 1); B) Within-CVI Characteristics score (Rating 2).

Figure 3 shows the changes in VBS scores over the same follow-up period. VBS scores improved (−0.32, p=0.008) in tandem with CVI Range-CR scores. Of note, the VBS is scored in the reverse direction as the CVI Range-CR, so the negative change here reflects improvement in functioning.

**Figure 3.**
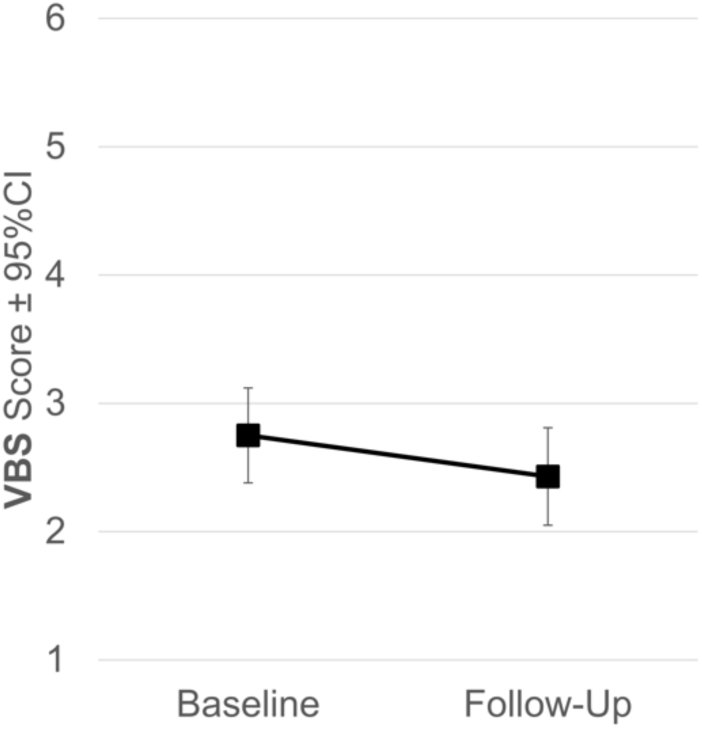
Change in Visual Behavior Scale (VBS) scores over 1 year.

## Discussion

In our cohort of 40 children with CVI, we report that the CVI Range-CR had excellent intra-rater reliability, good-to-excellent inter-rater reliability, and strong validity when compared to clinical assessment of visual behavior. Additionally, we found that the 10 domains used to calculate Rating 2 of the CVI Range-CR had strong internal consistency, supporting their combination into a single score.

For Rating 2 of the CVI Range-CR, Cronbach’s ɑ and McDonald’s ⍵ were both 0.96, indicating high internal consistency or relationship between the domains. Item-test correlation for all domains was greater than 0.80, except for Atypical Visual Reflexes, which was 0.69. This suggests that the visual reflex response may be assessing a different construct than the other domains of the CVI Range-CR. When evaluating visual reflexes, the examiner notes whether the child blinks in response to both a visual threat and touching the face. Thus, the “visual reflexes” assessed in the CVI Range-CR are dependent on both visual and tactile input. This likely accounts for the lower item-test correlation in the Atypical Visual Reflexes domain. However, when this domain was excluded, Cronbach’s ɑ was unchanged at 0.96. Therefore, although the Atypical Visual Reflexes domain is less correlated to the overall score and other domains within the CVI Range-CR, the data do not support the necessity of removing this domain from the scale. Confirmatory factor analysis also suggests that the scale does not require adjustment, since the RMSEA was low at 0.045 and the coefficient of determination was high at 0.965.

Both Ratings 1 and 2 demonstrated excellent intra-rater reliability (ICC 0.88-0.98) when remote graders scored the same videos at two different timepoints. However, intra-rater reliability for individual domains was lower, and in many cases the confidence intervals (CIs) were wide. For grader 2, the CIs crossed 0 for the following three domains: Visual Field Preferences, Atypical Visual Reflexes, and Difficulty with Visual Novelty. In the CVI Range-CR, rather than performing traditional confrontational visual field testing, the examiner projects a bright colored flashlight onto the child’s face from various peripheral locations and notes whether the child reacts to the light. It may be challenging to determine remotely whether the child sees the light from the periphery because the light source may in some cases be an aversive stimulus, leading the child to ignore or avoid the light instead of looking toward it. Subtle responses may be missed when reviewing the video recording due the angle or distance of the iPad from the child. As noted above, Atypical Visual Reflexes are graded based on blink to visual threat or touching the face. This may be difficult to assess on a video because the child’s eyes may be blocked by the examiner’s hand. Finally, Difficulty with Visual Novelty can only be assessed by history in the CVI Range-CR because we use a standardized set of materials which are all novel to the child. There may be some degree of subjective interpretation of historical information when this domain is graded, leading to imperfect intra-rater reliability.

Inter-rater reliability was also high for both Ratings 1 and 2 (absolute ICC 0.74 and 0.76, relative ICC 0.84 and 0.88). However, similar to intra-rater reliability, ICC scores were lower for individual domains, and confidence intervals were wide for many domains. The lowest inter-rater ICC values for absolute agreement were 0.46 (95% CI 0.24-0.68) for Difficulty with Visual Novelty and 0.48 (95% CI 0.24-0.72) for Need for Light. The potential challenges with assessing Difficulty with Visual Novelty in the CVI Range-CR are discussed above. For the Need for Light domain, the ICC improved from 0.48 to 0.62 when relative rather than absolute agreement was considered. Relative ICC values take into account static differences between graders. This suggests that the variability in the Need for Light domain is in part because the graders had different thresholds for determining whether a child required light to facilitate functional vision. This difference between absolute and relative ICC values was seen on several domains of Rating 2.

We assessed validity of CVI Range-CR scores by comparison to clinical assessment of visual acuity using the six-level Visual Behavior Scale (VBS). We used the VBS as a comparator because there is no gold standard to measure functional vision in children with CVI. Visual acuity is only one aspect of vision, and the CVI Range-CR assesses multiple domains of functional vision. However, visual discrimination, as measured by visual acuity, is a prerequisite for certain aspects of functional vision, such as the ability to identify objects on a crowded background. Therefore, measures of functional vision should correlate to some degree with measures of visual acuity. We found a significant and strong correlation between CVI Range-CR and VBS scores (correlation coefficient −0.76 to −0.86, p<0.0001), supporting the validity of the CVI Range-CR as a visual assessment.

We also correlated CVI Range-CR scores to adaptive function, as assessed by the VABS-III. VABS-III composite, domain, and subdomain scores have been shown to be significantly abnormal in children with CVI, with the average reported scores lower than the first percentile.^30^ Our previous study found that in children with CVI, weakly significant correlations were seen between VBS scores and the VABS-III composite, Socialization domain, and Interpersonal Relationships and Play and Leisure subdomains (correlation coefficient≥0.30). In the present study, the correlations between CVI Range-CR and VABS-III scores were significant but mostly weak, although they reached the moderate level in the Communication, Daily Living Skills, and Motor Skills domains, as well as the Interpersonal Relationships and Play and Leisure subdomains (correlation coefficient≥0.421). The stronger correlation between CVI Range-CR and VABS-III scores compared to VBS and VABS-III scores suggests that functional vision may be more important than visual acuity alone in the development of age-appropriate behaviors across multiple domains in children with CVI.

When CVI Range-CR assessments were compared over one year, there was on average a small improvement in functional vision (0.31 to 0.35 points). This difference is smaller than the difference in scores between graders (range 0.28 to 1.21). Therefore, it is recommended that future prospective, longitudinal studies of children with CVI use the same grader for CVI Range-CR assessments conducted at multiple time points. The VBS also showed a small improvement from 2.6 to 2.3 over 1 year, which further supports the validity of the CVI Range-CR for longitudinal assessments. Because the participants in our study generally did not receive targeted CVI interventions, the natural history data from this study may inform sample size calculations for future interventional studies. Our planned future research will examine factors associated with change in CVI Range-CR scores over 1 year in this cohort, such as treatment of medical conditions or various therapies. This will also inform future interventional studies.

The present study should be interpreted in light of its limitations. The data was skewed toward participants with lower levels of functional vision, and only six (15%) participants were in phase 3 (with CVI Range-CR scores greater than 7) at either the baseline or 1 year visit, based on scoring by the in-person examiner. This is largely due to the nature of patients in our tertiary referral clinic, who typically have more severe disease than those in the general population. We also had six (15%) participants who were lost to follow-up, potentially leading to bias in our 1 year follow-up results. However, the cohort of 34 participants that completed the 1 year follow-up visit was not significantly different from the six participants lost to follow-up based on demographics, VBS scores, and CVI Range-CR scores at baseline.

In summary, we report that scoring of recorded CVI Range-CR assessments is reliable and valid. This suggests that future multi-site studies could potentially record CVI Range-CR assessments for scoring by a centralized reading center, which would take advantage of the test’s excellent intra-rater reliability. Inter-rater reliability, while still considered good, was lower than intra-rater reliability, and the range of differences between graders was larger than the change in CVI Range-CR scores over 1 year. This suggests that consistent graders should be used if children with CVI undergo assessments at multiple time points. We acknowledge that there remains a significant time burden to training, administering, and scoring the test, which could be a barrier to including CVI Range-CR assessments in multi-center research studies. Planned future research will address these issues using artificial intelligence to develop a short version of the CVI Range-CR with automated scoring, facilitating the inclusion of functional vision assessments in future clinical trials for CVI. This is critical to ensure that evidence-based medical treatments for CVI improve not only visual function that clinicians measure in the eye clinic, but also a child’s ability to use their vision in activities of daily life at home and school.

## Data Availability

All data produced in the present study are available upon reasonable request to the authors.

## Other Acknowledgments

We wish to acknowledge Randy Nguyen, BS (University of Southern California) for assistance with editing videos.

