## Supplemental Data for "Validity and Reliability of the CVI Range for Clinical Research (CVI Range-CR): Baseline and One-Year Results"

Supplemental Table 1. Six-level Visual Behavior Scale (VBS).

| | |
|---|---|
| 6 | No visual behavior |
| 5 | Behavioral response to light |
| 4 | Behavioral response to motion |
| 3 | Poor fixation with ability to follow face or large object |
| 2 | Fixation and pursuit of a 6-inch toy at 1 foot |
| 1 | Fixation and pursuit of a 1-inch toy at 1 foot |

Supplemental Table 2. Inter-rater reliability of 3 graders (one in-person, two remote who alternated scoring) for 40 CVI Range-CR assessments at baseline only.

| | **Absolute Agreement** | | **Relative Agreement** | |
|---|---|---|---|---|
| | **ICC** | **95% CI** | **ICC** | **95% CI** |
| Rating 1 – Across-CVI characteristics | 0.76 | 0.59 – 0.92 | 0.84 | 0.75 – 0.90 |
| Rating 2 – Within-CVI characteristics | 0.76 | 0.53 – 0.99 | 0.88 | 0.81 – 0.92 |
| Rating 2 Domains | | | | |
| 1. Color Preference | 0.63 | 0.43 – 0.82 | 0.67 | 0.48 – 0.81 |
| 2. Need for Movement | 0.57 | 0.31 – 0.82 | 0.67 | 0.48 – 0.82 |
| 3. Visual Latency | 0.61 | 0.33 – 0.90 | 0.76 | 0.61 – 0.87 |
| 4. Visual Field Preferences | 0.46 | 0.21 – 0.71 | 0.51 | 0.29 – 0.72 |
| 5. Difficulties with Visual Complexity | 0.61 | 0.37 – 0.85 | 0.72 | 0.55 – 0.84 |
| 6. Need for Light | 0.40 | 0.15 – 0.66 | 0.47 | 0.25 – 0.70 |
| 7. Difficulty with Distance Viewing | 0.71 | 0.51 – 0.90 | 0.78 | 0.64 – 0.88 |
| 8. Atypical Visual Reflexes | 0.70 | 0.53 – 0.87 | 0.70 | 0.53 – 0.83 |
| 9. Difficulty with Visual Novelty | 0.52 | 0.23 – 0.81 | 0.66 | 0.47 – 0.81 |
| 10. Absence of Visually Guided Reach | 0.75 | 0.60 – 0.90 | 0.79 | 0.64 – 0.88 |

INTERNAL

Supplemental Table 3. Inter-rater reliability of 3 graders (one in-person, two remote) for 34 CVI Range-CR assessments at 1 year follow-up visit only.

| | **Absolute Agreement** | | **Relative Agreement** | |
|---|---|---|---|---|
| | **ICC** | **95% CI** | **ICC** | **95% CI** |
| Rating 1 – Across -CVI characteristics | 0.75 | 0.57 – 0.94 | 0.84 | 0.74 – 0.91 |
| Rating 2 – Within-CVI characteristics | 0.74 | 0.47 – 0.99 | 0.88 | 0.81 – 0.93 |
| Rating 2 Domains | | | | |
| 1. Color Preference | 0.61 | 0.40 – 0.83 | 0.70 | 0.55 – 0.82 |
| 2. Need for Movement | 0.60 | 0.40 – 0.79 | 0.66 | 0.49 – 0.79 |
| 3. Visual Latency | 0.65 | 0.42 – 0.88 | 0.76 | 0.63 – 0.86 |
| 4. Visual Field Preferences | 0.50 | 0.26 – 0.75 | 0.61 | 0.44 – 0.76 |
| 5. Difficulties with Visual Complexity | 0.53 | 0.28 – 0.78 | 0.64 | 0.47 – 0.78 |
| 6. Need for Light | 0.52 | 0.22 – 0.82 | 0.69 | 0.54 – 0.82 |
| 7. Difficulty with Distance Viewing | 0.64 | 0.46 – 0.83 | 0.70 | 0.54 – 0.82 |
| 8. Atypical Visual Reflexes | 0.58 | 0.34 – 0.83 | 0.70 | 0.54 – 0.82 |
| 9. Difficulty with Visual Novelty | 0.54 | 0.31 – 0.78 | 0.65 | 0.48 – 0.78 |
| 10. Absence of Visually Guided Reach* | 0.70 | 0.52 – 0.89 | 0.77 | 0.64 – 0.87 |

*Rated for only n=28 assessments at follow-up, reflecting participant functioning

Supplemental Table 4. Correlation between CVI Range-CR and Visual Behavior Scale (VBS) scores at baseline visit only.

| | **CVI Range-CR Rating 1** | | **CVI Range-CR Rating 2** | |
|---|---|---|---|---|
| | Correlation coefficient | p-value | Correlation coefficient | p-value |
| In-person grader | -0.84 | <0.0001 | -0.84 | <0.0001 |
| Remote grader 1 | -0.79 | <0.0001 | -0.76 | 0.0001 |
| Remote grader 2 | -0.75 | 0.0001 | -0.80 | <0.0001 |
| Average | -0.86 | <0.0001 | -0.86 | <0.0001 |

Supplemental Table 5. Correlation between CVI Range-CR and Visual Behavior Scale (VBS) scores at 1 year follow-up visit only.

| | **CVI Range-CR Rating 1** | | **CVI Range-CR Rating 2** | |
|---|---|---|---|---|
| | Correlation coefficient | p-value | Correlation coefficient | p-value |
| In-person grader | -0.84 | <0.0001 | -0.84 | <0.0001 |
| Remote grader 1 | -0.79 | <0.0001 | -0.79 | <0.0001 |
| Remote grader 2 | -0.80 | <0.0001 | -0.82 | <0.0001 |
| Average | -0.83 | <0.0001 | -0.84 | <0.0001 |

INTERNAL

Supplemental Table 6. Correlation between CVI Range-CR and Vineland Adaptive Behavior Scales, 3rd edition (VABS-III) scores, including baseline visits only. The average CVI Range-CR scores from two graders were used for this analysis.

| | **CVI Range-CR Rating 1** | | **CVI Range-CR Rating 2** | |
|---|---|---|---|---|
| | Correlation coefficient | p-value | Correlation coefficient | p-value |
| Adaptive Behavior Composite (ABC) | 0.375 | 0.024* | 0.409 | 0.013* |
| Communication | | | | |
| Domain | 0.391 | 0.018* | 0.421 | 0.010* |
| Subdomains | | | | |
| Receptive | 0.388 | 0.019* | 0.409 | 0.013* |
| Expressive | 0.373 | 0.025* | 0.410 | 0.013* |
| Written | 0.318 | 0.114 | 0.336 | 0.093 |
| Daily Living Skills | | | | |
| Domain | 0.289 | 0.088 | 0.316 | 0.060 |
| Subdomains | | | | |
| Personal | 0.433 | 0.008* | 0.462 | 0.005* |
| Domestic | 0.111 | 0.589 | 0.138 | 0.502 |
| Community | 0.281 | 0.165 | 0.292 | 0.148 |
| Socialization | | | | |
| Domain | 0.368 | 0.027* | 0.413 | 0.012* |
| Subdomains | | | | |
| Interpersonal Relationships | 0.451 | 0.006* | 0.495 | 0.002* |
| Play and Leisure | 0.400 | 0.016* | 0.449 | 0.006* |
| Coping Skills | 0.285 | 0.135 | 0.316 | 0.095 |
| Motor Skills | | | | |
| Domain | 0.514 | 0.002 | 0.540 | <0.001* |
| Subdomains | | | | |
| Gross Motor | 0.426 | 0.010* | 0.440 | 0.007* |
| Fine Motor | 0.461 | 0.005* | 0.494 | 0.002* |

Supplemental Table 7. Correlation between CVI Range-CR and Vineland Adaptive Behavior Scales, 3rd edition (VABS-III) scores, including 1 year follow-up visits only. The average CVI Range-CR scores from three graders were used for this analysis.

| | **CVI Range-CR Rating 1** | | **CVI Range-CR Rating 2** | |
|---|---|---|---|---|
| | Correlation coefficient | p-value | Correlation coefficient | p-value |
| Adaptive Behavior Composite (ABC) | 0.68 | <0.0001 | 0.65 | <0.0001 |
| Communication | | | | |
| Domain | 0.71 | <0.0001 | 0.70 | <0.0001 |
| Subdomains | | | | |
| Receptive | 0.40 | 0.02 | 0.41 | 0.02 |
| Expressive | 0.51 | 0.0027 | 0.51 | 0.0026 |
| Written | 0.59 | 0.0011 | 0.58 | 0.0016 |
| Daily Living Skills | | | | |
| Domain | 0.61 | 0.0002 | 0.57 | 0.0006 |
| Subdomains | | | | |
| Personal | 0.56 | 0.0009 | 0.57 | 0.0006 |
| Domestic | 0.53 | 0.0042 | 0.51 | 0.007 |
| Community | 0.59 | 0.0012 | 0.56 | 0.0025 |
| Socialization | | | | |
| Domain | 0.70 | <0.0001 | 0.67 | <0.0001 |
| Subdomains | | | | |
| Interpersonal Relationships | 0.73 | <0.0001 | 0.70 | <0.0001 |
| Play and Leisure | 0.71 | <0.0001 | 0.69 | <0.0001 |
| Coping Skills | 0.60 | 0.0003 | 0.57 | 0.0006 |
| Motor Skills | | | | |
| Domain | 0.77 | <0.0001 | 0.78 | <0.0001 |
| Subdomains | | | | |
| Gross Motor | 0.56 | 0.001 | 0.56 | 0.001 |
| Fine Motor | 0.64 | <0.0001 | 0.65 | <0.0001 |

### Supplemental Forms

#### Baseline Questionnaire: CVI Range-CR Study Intake Form

Child's Age: _________ Gender: ☐ Male ☐ Female
Person filling out form: Mother ☐ Father ☐ Legal Guardian ☐
Other_______________
What is the primary language spoken in the home? ______________
Other languages spoken in the home: ______________
Home Zip Code: _________

#### Family information and history

Parent 1

**Parent Age:** ___________________
**Living?** ☐ Yes ☐ No
**Occupation**: ______________________________
**Years of School completed**: ☐ Less than 8 ☐ Less than 12 ☐ High school diploma (12 yrs) ☐ Some college (13-15 yrs) ☐ Associate's Degree (14 yrs) ☐ Bachelor's degree (16 yrs) ☐ Some graduate school (> 16 yrs) ☐ Completed graduate degree (>16 yrs)
**Race**: ☐ African American/Black ☐ American Indian/Alaska Native ☐ Asian/Pacific Islander ☐ Caucasian ☐ Hispanic/Latino ☐ Multiracial ☐ Other: _______________________
**Ethnicity:** ☐ Hispanic/Latino ☐ Not Hispanic/Latino
**Marital Status**: ☐ Single ☐ Married ☐ Divorced/Separated ☐ Widowed ☐ Cohabitating
**How often does this child have contact with this parent?** ☐ Daily ☐ Weekly ☐ Bi-weekly ☐ Bi-monthly ☐ Monthly ☐ Several times a year ☐ Annually ☐ No contact

Parent 2

**Parent Age:** ___________________
**Living?** ☐ Yes ☐ No
**Occupation**: ______________________________
**Years of School completed**: ☐ Less than 8 ☐ Less than 12 ☐ High school diploma (12 yrs) ☐ Some college (13-15 yrs) ☐ Associate's Degree (14 yrs) ☐ Bachelor's degree (16 yrs) ☐ Some graduate school (> 16 yrs) ☐ Completed graduate degree (>16 yrs)
**Race**: ☐ African American/Black ☐ American Indian/Alaska Native ☐ Asian/Pacific Islander ☐ Caucasian ☐ Hispanic/Latino ☐ Multiracial ☐ Other: _______________________
**Ethnicity:** ☐ Hispanic/Latino ☐ Not Hispanic/Latino
**Marital Status**: ☐ Single ☐ Married ☐ Divorced/Separated ☐ Widowed ☐ Cohabitating
**How often does this child have contact with this parent?** ☐ Daily ☐ Weekly ☐ Bi-weekly ☐ Bi-monthly ☐ Monthly ☐ Several times a year ☐ Annually ☐ No contact

What is the approximate combined annual family income? ☐ less than $19,999 ☐ $20,000 to $39,999 ☐ $40,000-$59,999 ☐ $60,000-$79,999 ☐ $80,000-$99,999 ☐ $100,000 to $149,999 ☐ $150,000 or above
How many children live in the household? ___________

How many people in total, including children, live in the household? __________

Please check if any family member (e.g., parent, sibling, 1st cousin) has been diagnosed with the following conditions:

- ☐ Cortical visual impairment (CVI)
- ☐ Other childhood visual disorder (please specify): ______________________
- ☐ Neurological Disorder
- ☐ Genetic Disorder

If you checked any boxes, please specify (e.g., which family member(s), age, treatment):

________________________________________________________________________________

__________________________________________________

_________________________________________________________________

### Pregnancy History

Did the mother receive prenatal care? ☐ Yes ☐ No

☐ Full Term (37-42 weeks) ☐ Premature _____ weeks
Birth weight: ______ lbs. ______ ozs.
Type of delivery: ☐ vaginal ☐ C-section

Any complications during pregnancy? ☐ yes ☐ no
Any complications during delivery? ☐ yes ☐ no
Any complications following delivery? ☐ yes ☐ no
Any additional pregnancy history (e.g., illness, accidents, major stresses)?

_________________________________________________________________

Was there a stay in the NICU/PICU? How long? _______________________________
Was there any treatment following birth?

_________________________________________________________________

Age of mother at birth of this child: ______
Age of father at birth of this child: ______

Did mother use any of the following during pregnancy:

| | | |
|---|---|---|
| Alcohol? | ☐ yes | ☐ no |
| Cigarettes? | ☐ yes | ☐ no |
| Other drugs? | ☐ yes | ☐ no |

### Developmental assessments and services

Has your child ever had a CVI Range assessment? Y / N

If so, who administered the test and when did this occur?

_______________________________________________

Have you previously heard about the CVI Range before you were invited to take part in this study? Y / N

If so, where did you learn about the CVI Range?

_______________________________________________

Are you familiar with the 10 visual characteristics of children with CVI that are included in the CVI Range? Y/ N

What types of services does your child receive?

a. Physical therapy
b. Occupational therapy
c. Speech therapy
d. Vision services
e. Other (please explain) _______________

How frequently does your child receive the above services?

_____________________________________________________________________________

### Medical history

**1.** Does your child have any of the following diagnoses? (Please circle/mark all that apply)

- ☐ Stroke or lack of oxygen to brain
- ☐ Periventricular leukomalacia
- ☐ Head trauma
- ☐ Seizures
- ☐ Hydrocephalus
- ☐ Brain abnormality on MRI (examples: schizencephaly, colpocephaly, porencephaly)
  Please specify: _______________________________________________________
- ☐ Meningitis, encephalitis, or meningoencephalitis
- ☐ Genetic disorder (please explain) ________________________________________
- ☐ Cerebral palsy

☐ Other neurologic condition(s) ________________________________________

2. Please list any other medical diagnoses:

________________________________________________________________________

________________________________________________________________________

________________________________________________________________________

________________________________________________________________________

3. Please list your child's medications:

________________________________________________________________________

________________________________________________________________________

________________________________________________________________________

________________________________________________________________________

4. Does your child have any of the following eye conditions? (Circle/mark all that apply)
   - ☐ Strabismus (eye misalignment)
   - ☐ Amblyopia ("lazy eye")
   - ☐ Refractive error (requires glasses)
   - ☐ Optic atrophy
   - ☐ Retinopathy of prematurity (ROP)

5. Please list any other eye conditions:

________________________________________________________________________

________________________________________________________________________

________________________________________________________________________

________________________________________________________________________

**Thank you for taking the time to fill out this questionnaire. We appreciate your time and effort.**

### CVI Range-CR Study 12-month Follow-Up Questionnaire

#### Developmental assessments and services

1. Did your child receive vision support services during the last 12 months?
   Yes / No

   If *yes*, proceed to question 2. If *no*, proceed to question 7.

2. Who provided the vision services? (Please circle)
   a. LAUSD
   b. Another school district (name): ________________________________
   c. Regional Center
   d. Other (explain): ____________________________________________

3. On average, how frequently did your child receive vision services over the past 12 months?
   a. Daily
   b. 4-5 times per week
   c. 1-3 times per week
   d. Other (explain): ____________________________________________

4. Did your child have a Teacher for the Visually Impaired (TVI)? Y / N
   a. Please provide the name of your child's TVI ______________________

5. Did your child receive CVI Range targeted intervention or support?
   a. Yes
   b. No
   c. Don't know

6. What types of vision interventions did your child receive?

   ___________________________________________________________

   ___________________________________________________________

   ___________________________________________________________

_______________________________________________

7. If your child did not receive vision services, what was the reason? (circle all that apply)
   a. Not part of local school district
   b. Did not meet age criteria for Regional Center services
   c. Did not qualify for vision therapy services based on school district or Regional Center assessment
   d. Other (explain): ___________________________________

7. In the last 12 months, has any family member (e.g., parent, sibling, 1st cousin) has been diagnosed with the following conditions:

- ☐ Cortical visual impairment (CVI)
- ☐ Other childhood visual disorder (please specify):
  _____________________________________________
- ☐ Neurological Disorder
- ☐ Genetic Disorder

If you checked any boxes, please specify (e.g., which family member(s), age, treatment):

______________________________________________________________

______________________________________

---

**Please complete the following if you have moved, changed jobs, or experienced other changes in your family's circumstances in the last 12 months.**

Parent 1

| |
|---|
| **Marital Status**: ☐ Single ☐ Married ☐ Divorced/Separated ☐ Widowed ☐ Cohabitating<br>**How often does this child have contact with this parent?** ☐ Daily ☐ Weekly ☐ Every other week ☐ Monthly ☐ Several times a year ☐ Annually ☐ No contact |

Parent 2

**Marital Status**: ☐ Single ☐ Married ☐ Divorced/Separated ☐ Widowed ☐ Cohabitating
**How often does this child have contact with this parent?** ☐ Daily ☐ Weekly
☐ Every other week ☐ Monthly ☐ Several times a year ☐ Annually ☐ No contact

What is your home zip code? ___________

What is the approximate combined annual family income? ☐ less than $19,999
☐ $20,000 to $39,999 ☐ $40,000-$59,999 ☐ $60,000-$79,999
☐ $80,000-$99,999 ☐ $100,000 to $149,999 ☐ $150,000 or above

How many children live in the household? ___________

How many people in total, including children, live in the household? ___________

### Medical history

1. Has your child been diagnosed with any new medical conditions in the last 12 months (since the last study visit)?
    a. Yes
    b. No

2. If yes to #1, please list the new medical diagnoses:

______________________________________________

______________________________________________

______________________________________________

______________________________________________

______________________________________________

3. Have your child's medications been changed in the last 12 months?
    a. Yes
    b. No

4. If yes to #3, please list your child's current medications:

______________________________________________

______________________________________________

5. Has your child been diagnosed with a new eye condition in the last 12 months?
    a. Yes
    b. No

6. If yes to #5, please list the new eye conditions:

### Scoring worksheets for Across- and Within-CVI Characteristics methods of scoring the CVI Range-CR (Ratings 1 and 2).

#### CVI Range-CR Scoring: Across-CVI Characteristics Assessment Method (Rating 1)

Rate the following statements as related to the student/child's visual behaviors by marking the appropriate column to indicate the methods used to support the scores:

***O*** = Information obtained through observation of the student/child
***I*** = Information obtained through interview regarding the student/child
***D*** = Information obtained through direct contact with the student/child

In the remaining columns, rate each statement with one of the following descriptors:

**R** = Represents a visual behavior that is resolving or approaching typical behavior
\+ = Describes current functioning of student/child
+/– = Partially describes the student/child emerging
– = Does not apply to student/child

**CVI Range 1-2: Student functions with minimal visual responses**

| **O** | **I** | **D** | R | + | +/- | - | |
|---|---|---|---|---|---|---|---|
| | | | | | | | May localize, but no appropriate fixations on objects or faces |
| | | | | | | | Consistently attentive to lights or perhaps ceiling fans |
| | | | | | | | Prolonged periods of latency in visual tasks |
| | | | | | | | Responds only in strictly controlled environments |
| | | | | | | | Objects viewed are a single color |
| | | | | | | | Objects viewed have movement and/or shiny or reflective properties |
| | | | | | | | Visually attends in near space only |
| | | | | | | | No blink in response to touch or visual threat |
| | | | | | | | No regard of the human face |

**CVI Range 3-4: Student functions with more consistent visual response**

| **O** | **I** | **D** | R | + | +/- | - | |
|---|---|---|---|---|---|---|---|
| | | | | | | | Visually fixates when the environment is controlled |
| | | | | | | | Less attracted to lights: can be redirected |
| | | | | | | | Latency slightly decreases after periods of consistent viewing |
| | | | | | | | May look at novel objects if they share characteristics of familiar objects |
| | | | | | | | Blinks in response to touch and/or visual threat, but the responses may be latent and/or inconsistent |
| | | | | | | | Has “favorite” color |
| | | | | | | | Shows strong visual field preferences |
| | | | | | | | May notice moving objects at 2 to 3 feet |
| | | | | | | | Look and touch completed as separate events |

**CVI Range 5-6: Student uses vision for functional tasks**

| **O** | **I** | **D** | R | + | +/- | - | |
|---|---|---|---|---|---|---|---|
| | | | | | | | Objects viewed may have two to three colors |
| | | | | | | | Light is no longer a distractor |
| | | | | | | | Latency present only when the student is tired, stressed, or overstimulated |
| | | | | | | | Movement continues to be an important factor for visual attention |
| | | | | | | | Student tolerates low levels of background noise |
| | | | | | | | Blink response to touch is consistently present |
| | | | | | | | Blink response to visual threat is intermittently present |
| | | | | | | | Visual attention now extends beyond near space, up to 4 to 6 feet |
| | | | | | | | May regard familiar faces when voices do not compete |

**CVI Range 7-8: Student demonstrates visual curiosity**

| **O** | **I** | **D** | R | + | +/- | - | |
|---|---|---|---|---|---|---|---|
| | | | | | | | Selection of toys or objects is less restricted; requires one to two sessions of "warm up" |
| | | | | | | | Competing auditory stimuli tolerated during periods of viewing; the student may now maintain visual attention on objects that produce music |
| | | | | | | | Blink response to visual threat consistently present |
| | | | | | | | Latency rarely present |
| | | | | | | | Visual attention extends to 10 feet with targets that produce movement |
| | | | | | | | Movement not required for attention at near distance |
| | | | | | | | Smiles at/regards familiar and new faces |
| | | | | | | | May enjoy regarding self in mirror |
| | | | | | | | Most high-contrast colors and/or familiar patterns regarded and interpreted |
| | | | | | | | Simple books, picture cards, or symbols regarded and interpreted |

**CVI Range 9-10: Student spontaneously uses vision for most functional activities at level approaching near typical**

| O | I | D | R | + | +/- | - | |
|---|---|---|---|---|---|---|---|
| | | | | | | | Selection of toys or objects not restricted to the familiar; visually curious in new settings |
| | | | | | | | Only the most complex environments affect visual response |
| | | | | | | | Latency never present |
| | | | | | | | No color or pattern preference |
| | | | | | | | Visual attention and interpretation extends beyond 20 feet |
| | | | | | | | Views and interprets information from non-backlit two-dimensional materials and simple images |
| | | | | | | | Uses vision to imitate actions |
| | | | | | | | Demonstrates memory of visual events |
| | | | | | | | Displays typical visual-social responses |
| | | | | | | | Visual fields unrestricted |
| | | | | | | | Look and reach completed as a single action |
| | | | | | | | Views and interprets information from non-backlit two-dimensional images presented on complex, visually dense backgrounds |

### CVI Range-CR Scoring: Within-CVI Characteristics Assessment Method (Rating 2)

Determine the level of CVI present or resolved in the 10 categories below and add to obtain total score. Rate the following CVI categories as related to the student/child's visual behaviors by circling the appropriate number (the CVI Progress Chart may be useful as a scoring guide):

**0** Full effect of the characteristic is present

**.25** Behavior on this characteristic has begun to change or improve

**.5** The characteristic is affecting visual functioning approximately half the time

**.75** Occasional effect of the characteristic; response is nearly like that of individuals the same age

**1** Resolving, approaching typical, or response is the same as others of the same age

| | | | | | |
|---|---|---|---|---|---|
| **1. Color Preference** | **0** | **.25** | **.5** | **.75** | **1** |
| **2. Need for movement** | **0** | **.25** | **.5** | **.75** | **1** |
| **3. Visual latency** | **0** | **.25** | **.5** | **.75** | **1** |
| **4. Visual field preferences** | **0** | **.25** | **.5** | **.75** | **1** |
| **5. Difficulties with visual complexity- object<br>array<br>sensory<br>faces** | **0<br>0<br>0<br>0<br>0** | **.25<br>.25<br>.25<br>.25<br>.25** | **.5<br>.5<br>.5<br>.5<br>.5** | **.75<br>.75<br>.75<br>.75<br>.75** | **1<br>1<br>1<br>1<br>1** |
| **6. Need for light** | **0** | **.25** | **.5** | **.75** | **1** |
| **7. Difficulty with distance viewing** | **0** | **.25** | **.5** | **.75** | **1** |

| | | | | | |
|---|---|---|---|---|---|
| **8. Atypical visual reflexes** | **0** | **.25** | **.5** | **.75** | **1** |
| **9. Difficulty with visual novelty** | **0** | **.25** | **.5** | **.75** | **1** |
| **10. Absence of visually guided reach** | **0** | **.25** | **.5** | **.75** | **1** |
